# Relationship between sexting and self-esteem, depression, anxiety, and stress among young people

**DOI:** 10.1101/2023.02.01.23285354

**Authors:** Petros Galanis, Zoe Katsiroumpa, Aglaia Katsiroumpa, Anastasia Tsakalaki, Stefanos Vasilopoulos

## Abstract

**Introduction:** The huge spread of the internet and especially of social media has led to new ways of communication, even erotic communication, especially among young people, replacing, in many cases, activities that until now required the face-to-face meeting of individuals.

**Aim:** To investigate the relationship between sexting and self-esteem, depression, anxiety, and stress among young people. Also, we investigated the impact of demographic characteristics on sexting.

**Methods:** A cross-sectional study was conducted with 368 young people aged 18-30 years old. We created an anonymous form of the study questionnaire with Google forms and we disseminated it through social media. Thus, we obtained a convenience sample. We used valid scales to measure sexting, self-esteem, depression, anxiety and stress among young people. All scales in our study had very good Cronbach ‘s alpha.

**Results:** Mean age of the participants was 23.7 years, while 82.3% was females. Mean every day time that participants spent on social media/platforms/applications was 4.7 hours. Participants used more often to communicate with others Instagram (86.8%) and Facebook (62.8%), and then Viber (29.9%), TikTok (9.8%), Snapchat (6%) and WhatsApp (5.1%). Frequency of texting was low among participants, while self-esteem level was average. Moreover, participants had higher levels of stress than depression and anxiety. We found that increased sexting was associated with decreased self-esteem (r=-0.3, p=0.02), increased depression (r=0.4, p=0.001), increased anxiety (r=0.3, p=0.005), and increased stress (r=0.4, p<0.001). Multivariable linear regression analysis identified that increased number of accounts on social media/platforms/applications (coefficient beta=0.07, 95% confidence interval=0.01 to 0.13, p-value=0.023) and increased age (coefficient beta=0.08, 95% confidence interval=0.01 to 0.13, p-value=0.003) was associated with increased sexting.

**Conclusions:** Understanding the relationship between sexting and self-esteem, depression, anxiety, and stress in young people will give policy makers the opportunity to develop appropriate health education programmes to reduce risky sexual behaviors.

## Introduction

The huge spread of the internet and especially of social media has led to new ways of communication, even erotic communication, especially among young people, replacing, in many cases, activities that until now required the face-to-face meeting of individuals. This changing context of communication has also brought a change in behaviors, which is where sexting emerges from (Walker et al., 2011). The prevalence of sexting among young people is showing an increasing trend with a very wide variation from 0.9% to 78% (Barrense-Dias et al., 2017). The large variations in the prevalence of sexting are due to different measurement tools and different study designs (Dake et al., 2012; Klettke et al., 2018; Temple et al., 2014).

Sexting among young people is becoming increasingly widespread, becoming an extremely important issue that needs systematic investigation, mainly because of the negative consequences it brings, especially when its content is republished or shared, leading to humiliation, harassment or insult (Barrense-Dias et al, 2017). Sexting is essentially defined as sending and receiving personal material with sexual content via mobile phones, social media, websites and platforms (Krieger, 2017).

The exposure of young people ‘s personal information on social media is constantly increasing. Thus, the young person is represented by his or her digital analogue, socialized and sexualized online. Particularly for young people, who by definition are in the phase of searching and developing their sexuality and sexual identity, it makes sense that the frequency of sexting is increased (Barrense-Dias et al., 2017). Sexting among young people is a predictor of sexual behaviors and may be associated with mental health disorders and risky behaviors (Baumgartner et al., 2012). The role of gender in sexting is controversial, as in some studies the prevalence of sexting is higher in boys and in others it is higher in girls (Baumgartner et al., 2012; Kerstens & Stol, 2014; Livingstone & Görzig, 2014; Vanden Abeele et al., 2012).

The negative consequences of sexting in young people are numerous and can be summarized in four broad categories (Doyle et al, 2021): (a) psychological consequences (victimisation, sexual bullying, mental health disorders, quality of life and emotional disturbances; (b) behavioural consequences (sexual activity and risky behaviours such as aggression); (c) consequences on young people ‘s relationships with others with these relationships being friends, family or personal; and (d) consequences on young people ‘s personal lives by sharing/exposing their personal moments to third parties.

To the best of our knowledge, there is no study in Greece that has investigated the relationship between sexting and self-esteem, depression, anxiety, and stress among young people. Thus, the aim of our study was to investigate the relationship between sexting and self-esteem, depression, anxiety, and stress among young people. Also, we investigated the impact of demographic characteristics on sexting.

## Methods

### Study design

A cross-sectional study was conducted with 368 young people aged 18-30 years old. We created an anonymous form of the study questionnaire with Google forms and we disseminated it through social media. Data collection was performed in September 2022. Thus, we obtained a convenience sample. The participants were informed about the purpose and methodology of the study so that they could decide whether or not they wished to participate in the study. The participants were then asked to complete the questionnaire without giving their details (full name), which preserved their anonymity. There was no time limit, so that participants were not pressed for time and did not lead to rushed responses. Each questionnaire was placed in a special opaque envelope to which only the research team had access. This was the best way to ensure (a) the informed consent of the participants to participate in the study, (b) the anonymity of the participants and (c) the confidentiality of the information to which only the research team had access. Study protocol was approved by the Ethics Committee of Faculty of Nursing of National and Kapodistrian University of Athens (reference number: 418, 22 September 2022).

### Measures

We collected the following demographic characteristics of the participants: gender, age, kind of relationship, educational level of mothers and fathers, residence during the adolescence, accounts on Facebook, Instagram, TikTok, Snapchat, Viber, WhatsApp, hours spending on Facebook, Instagram, TikTok, Snapchat, Viber, WhatsApp, and communication with others through Facebook, Instagram, TikTok, Snapchat, Viber, WhatsApp.

To assess sexting among participants, the questionnaire of Bianchi et al. (2018) was used, which consists of 13 items (Bianchi et al., 2018). Each item of the questionnaire as well as the total score is given values from 1 to 5. Higher values in the questionnaire indicate a higher frequency of sexting. The questionnaire is reliable and valid in Greek (Galanis et al., 2022). In our study, Cronbach ‘s alpha for the questionnaire was 0.81.

The “Rosenberg Self-Esteem scale” was used to assess self-esteem in participants, which consists of 10 items and receives a total score from 10 to 40 (Rosenberg, 1965). An increase in the score indicates higher self-esteem. Validation of the “Rosenberg Self-Esteem scale” has already performed in Greek (Galanou et al. 2014). In our study, Cronbach ‘s alpha for the “Rosenberg Self-Esteem scale” was 0.86.

The “Depression Anxiety Stress Scales” was used to assess depression, anxiety and stress in participants (Lovibond & Lovibond 1995). Depression, anxiety and stress are measured by seven items each one and take values from 0 to 42. An increase in score indicates more depression, anxiety and stress. The “Depression Anxiety Stress Scales” are proven reliable and valid in Greek (Lyrakos et al. 2011). In our study, Cronbach ‘s alpha for the depression, anxiety and stress scales was 0.86, 0.78, and 0.89 respectively.

### Statistical analysis

We present categorical variables with numbers (percentages) and continuous variables with mean (standard deviation). We used the Kolmogorov-Smirnov test to assess the distribution of the continuous variables. We found that the continuous variables followed the normal distribution. Bivariate analysis included independent samples t-test, analysis of variance, Pearson ‘s correlation coefficient, and Spearman ‘s correlation coefficient. We used multivariable linear regression analysis to assess the impact of demographic characteristics on sexting. All tests of statistical significance were two-tailed, and p-values<0.05 were considered as statistically significant. Statistical analysis was performed with the IBM SPSS 21.0 (IBM Corp. Released 2012. IBM SPSS Statistics for Windows, Version 21.0 Armonk, NY, USA).

## Results

Study sample included 368 young people aged 18-30 years old. Demographic characteristics of the participants are shown in Table 1. Mean age of the participants was 23.7 years. Among the participants, 82.3% was females, 41.3% was in a serious relationship, and 50.3% were living in an urban area during the adolescence.

**Table 1.** Demographic characteristics of the 368 participants.

| Characteristics | N | % |
| --- | --- | --- |
| Gender |  |  |
| Females | 303 | 82.3 |
| Males | 65 | 17.7 |
| Age (mean, standard deviation) | 23.7 | 2.5 |
| Relationship |  |  |
| No | 184 | 49.9 |
| Open | 32 | 8.8 |
| Serious | 152 | 41.3 |
| Mother's educational level |  |  |
| Elementary school | 25 | 6.9 |
| High school | 160 | 43.5 |
| University degree | 183 | 49.6 |
| Father's educational level |  |  |
| Elementary school | 43 | 11.8 |
| High school | 189 | 51.3 |
| University degree | 136 | 36.9 |
| Residence during the adolescence |  |  |
| Urban area | 185 | 50.3 |
| Semi urban area | 138 | 37.5 |
| Rural area | 35 | 12.2 |

Social media/platforms/applications use of the participants is shown in Table 2. Mean every day time that participants spent on social media/platforms/applications was 4.7 hours (2.9), while minimum value was 0.1 hours and maximum value was 16 hours. Participants used more often to communicate with others Instagram (86.8%) and Facebook (62.8%), and then Viber (29.9%), TikTok (9.8%), Snapchat (6%) and WhatsApp (5.1%).

**Table 2.** Social media/platforms/applications use of the participants.

|  | Facebook | Instagram | TikTok | Snapchat | Viber | WhatsApp |
| --- | --- | --- | --- | --- | --- | --- |
| Account on | 191 (81.6) | 217 (92.7) | 153 (65.4) | 74 (31.6) | 179 (76.5) | 37 (15.8) |
| Communication with others through | 147 (62.8) | 203 (86.8) | 23 (9.8) | 14 (6) | 70 (29.9) | 12 (5.1) |

Values are expressed as numbers (percentages).

Descriptive statistics for sexting, self-esteem, depression, anxiety, and stress scales are shown in Table 3. Frequency of texting was low among participants, while self-esteem level was average. Moreover, participants had higher levels of stress than depression and anxiety.

**Table 3.** Descriptive statistics for sexting, self-esteem, depression, anxiety, and stress scales.

| Scale | Mean | Standard deviation | Minimum value | Maximum value |
| --- | --- | --- | --- | --- |
| Sexting | 1.7 | 0.8 | 1 | 3.5 |
| Self-esteem | 28.2 | 4.1 | 16 | 38 |
| Depression | 10.1 | 9.3 | 0 | 42 |
| Anxiety | 8.9 | 8.2 | 0 | 36 |
| Stress | 14.9 | 9.5 | 0 | 42 |

Relationships between sexting and self-esteem, depression, anxiety, and stress scales are shown in Table 4. We found that increased sexting was associated with decreased self-esteem (r=-0.3, p=0.02), increased depression (r=0.4, p=0.001), increased anxiety (r=0.3, p=0.005), and increased stress (r=0.4, p<0.001).

**Table 4.** Relationships between sexting and self-esteem, depression, anxiety, and stress scales.

|  | Sexting |  |
| --- | --- | --- |
|  | Pearson's correlation coefficient | P-value |
| Self-esteem | -0.3 | 0.02 |
| Depression | 0.4 | 0.001 |
| Anxiety | 0.3 | 0.005 |
| Stress | 0.4 | <0.001 |

**Table 5.** Bivariate analysis between demographic characteristics of the participants and sexting score.

| Independent variables | Mean sexting score | Standard deviation | P-value |
| --- | --- | --- | --- |
| Gender |  |  | 0.12<sup>a</sup> |
| Females | 1.73 | 0.59 |  |
| Males | 1.51 | 0.61 |  |
| Age |  | 0.3<sup>b</sup> | 0.01<sup>b</sup> |
| Mother's educational level |  | 0.027<sup>c</sup> | 0.7<sup>c</sup> |
| Father's educational level |  | -0.12<sup>c</sup> | 0.1<sup>c</sup> |
| Relationship |  |  | 0.3<sup>d</sup> |
| No | 1.69 | 0.62 |  |
| Open | 1.71 | 0.63 |  |
| Serious | 1.73 | 0.61 |  |
| Residence during the adolescence |  |  | 0.9<sup>d</sup> |
| Urban area | 1.61 | 0.62 |  |
| Semi urban area | 1.62 | 0.65 |  |
| Rural area | 1.56 | 0.66 |  |
| Every day hours that participants spent on social media/platforms/applications |  | 0.12<sup>b</sup> | 0.22<sup>b</sup> |
| Number of accounts on social media/platforms/applications |  | 0.11<sup>c</sup> | 0.04<sup>c</sup> |
| Number social media/platforms/applications that participants use to communicate with others |  | 0.16<sup>c</sup> | 0.11<sup>c</sup> |

<sup>a</sup> independent samples t-test
<sup>b</sup> Pearson's correlation coefficient
<sup>c</sup> Spearman's correlation coefficient
<sup>d</sup> analysis of variance

Bivariate analysis between demographic characteristics of the participants and sexting score are shown in Table 2. Multivariable linear regression analysis identified that increased number of accounts on social media/platforms/applications (coefficient beta=0.07, 95% confidence interval=0.01 to 0.13, p-value=0.023) and increased age (coefficient beta=0.08, 95% confidence interval=0.01 to 0.13, p-value=0.003) was associated with increased sexting.

## Discussion

We conducted a cross-sectional study to investigate the relationship between sexting and self-esteem, depression, anxiety, and stress among young people.

We found that lower self-esteem was associated with an increased frequency of sexting among young people, a finding that is confirmed by studies in other countries such as Germany, Spain, Australia, the USA and Croatia (Eugene, 2015; Klettke et al., 2019; Schoeps et al., 2020; Scholes-Balog et al., 2016; Sesar, 2021; Tamarit et al., 2021; Wachs et al., 2017). One possible explanation for this relationship is that young people who have a negative self-image and underestimate their abilities tend to doubt themselves. For this reason, they try to increase their self-confidence by seeking both communication with other people and validation from others. It is also typical that young people with low self-esteem also have a worse body image which leads them sexting more often (Frost & McKelvie, 2004).

In addition, we found that increased depression, anxiety and stress in young people were associated with increased frequency of sexting. This finding is supported by several studies that have found sexting to be associated with mental health problems in young people such as depression, anxiety and stress (Chaudhary et al., 2017; Dake et al., 2012; Eugene, 2015; Frankel et al., 2018; Gámez-Guadix & de Santisteban, 2018; Kim et al., 2020; Klettke et al., 2018, 2019; Temple et al., 2014; Van Ouytsel et al., 2014; Ybarra & Mitchell, 2014). The amount of time that young people spend on the internet and especially on social media has increased significantly in recent years, particularly with the emergence of smart phones that provide easy and quick access to social media. As a result, the internet is now one of the most common ways for young people to communicate with each other. This communication includes communication on an erotic level, including sexting. However, young people are not yet mature enough to manage the internet use properly, so that in many cases they spend an excessive number of hours on it and even engage in risky sexual behavior at the same time. As a result, the increased use of social media, and more specifically sexting, leads young people to mental health problems such as depression, anxiety and stress. Particular attention needs to be paid to this link between sexting and mental health problems in young people as if not addressed early and effectively they can lead to even worse problems such as aggressive behavior, substance use, suicidal tendencies etc. For this reason, parents and teachers need to identify dangerous behavior of young people online, so that any mental health problems can be managed more effectively.

Regarding the effect of demographic characteristics on the frequency of sexting, we found that older teenagers sexting more often. This finding is confirmed by similar studies (Gámez-Guadix & de Santisteban, 2018; Molla-Esparza et al., 2021; Ybarra & Mitchell, 2014). It seems that as they get older, people have less hesitations and reservations regarding the use of sexting. In addition, older people may want to experiment more about their sex lives, with such experimentation including sexting. We also found that young people with more social media accounts sexting more frequently. This finding makes sense, as young people with more social media accounts are also quite likely to use these media more which increases the likelihood that they will use them for sexting.

The present study also had some limitations. Specifically, the information on the young people was obtained at a specific point in time without knowing the temporal sequence between the variables. For this reason, there is a need for studies that should follow the young people for a period of time in order to draw more secure conclusions about the relationship between sexting and self-esteem, depression, anxiety, and stress. In addition, the study was conducted on a specific sample of young people. Therefore, the results cannot be generalized to all young people in Greece. Therefore, further studies with different study samples need to be conducted in order to draw more valid conclusions. The questionnaires to assess sexting, self-esteem, depression, anxiety and stress in the young people were self-completed. Finally, it is possible that there are other factors associated with sexting such as personality traits, psychological factors, etc. It is clear that more and larger sample size studies are needed in order to address errors to the greatest extent possible and draw safer conclusions.

In conclusion, although the present study found relationships between sexting and self-esteem, depression, anxiety and stress in young people, it is necessary to conduct further studies on other samples of young people in order to increase our knowledge in this research field. A better understanding of the relationship between sexting and self-esteem, depression, anxiety and stress in young people will also enable the development of appropriate health and internet safety education programmes to reduce risky sexual behaviors. Parents, teachers and healthcare professionals should pay particular attention on sexting. For example, healthcare professionals could ask young people about their sexual behavior in order to identify those who engage in risky sexual behaviors and provide them with appropriate information. Parents and teachers, moreover, need more and better education to understand the negative effects of sexting on the health of young people.

## Data Availability

All data produced in the present study are available upon reasonable request to the authors

## Notes

### Competing Interest Statement

The authors have declared no competing interest.

### Funding Statement

This study did not receive any funding

### Author Declarations

Study was approved by the Ethics Committee of Faculty of Nursing of National and Kapodistrian University of Athens (reference number: 418, 22 September 2022).

